# Distinct White Matter Fiber Density Patterns in Bipolar and Depressive Disorders: Insights from Fixel-Based Analysis

**DOI:** 10.1101/2025.02.19.25322569

**Authors:** Anna Manelis, Hang Hu, Skye Satz, Satish Iyengar, Holly A. Swartz

## Abstract

**Background:** Differentiating Bipolar (BD) and depressive (DD) disorders remains challenging in clinical practice due to overlapping symptoms. Our study employs fixel-based analysis (FBA) to examine fiber-specific white matter differences in BD and DD and gain insights into the ability of FBA metrics to predict future spectrum mood symptoms.

**Methods:** 163 individuals between 18 and 45 years with BD, DD, and healthy controls (HC) underwent Diffusion Magnetic Resonance Imaging. FBA was used to assess fiber density (FD), fiber cross-section (FC), and fiber density cross-section (FDC) in major white matter tracts. A longitudinal follow-up evaluated whether FBA measures predicted future spectrum depressive and hypomanic symptom trajectories over six months.

**Results:** Direct comparisons between BD and DD indicated lower FD in the right superior longitudinal and uncinate fasciculi and left thalamo-occipital tract in BD versus DD. Individuals with DD exhibited lower FD in the left arcuate fasciculus than those with BD. Compared to HC, both groups showed lower FD in the splenium of the corpus callosum and left striato-occipital and optic radiation tracts. FD in these tracts predicted future spectrum symptom severity. Exploratory analyses revealed associations between FD, medication use, and marijuana exposure.

**Conclusions:** Our findings highlight distinct and overlapping white matter alterations in BD and DD. Furthermore, FD in key tracts may serve as a predictor of future symptom trajectories, supporting the potential clinical utility of FD as a biomarker for mood disorder prognosis. Future longitudinal studies are needed to explore the impact of treatment and disease progression on white matter microstructure.

## INTRODUCTION

Bipolar (BD) and depressive (DD) disorders are severe mental illnesses that affect psychosocial functioning, reduce quality of life, and increase the risk of premature mortality in millions globally (Ferrari, 2022; Greenberg et al., 2021; Gutiérrez-Rojas et al., 2020; McIntyre et al., 2020). DD is characterized by episodes of depression (Greenberg et al., 2021), while BD is characterized by both depressive and hypo/manic episodes (McIntyre et al., 2020). Despite distinct clinical manifestations, accurately distinguishing between the two disorders remains challenging in clinical practice, leading to initial misclassification of bipolar disorder in at least 60% of cases and an average treatment delay of at least 5 years (Dagani et al., 2017; Drancourt et al., 2013; Hirschfeld et al., 2003; McIntyre et al., 2020). Given the substantial impact of diagnostic uncertainty on long-term outcomes, a deeper understanding of the neurobiological differences between BD and DD is warranted (Han et al., 2019; Phillips and Swartz, 2014).

Diffusion Magnetic Resonance Imaging (dMRI) is a neuroimaging method used to investigate the properties of white matter tracts by analyzing the movement of water molecules within them (Jones et al., 2013; Yeh et al., 2021). Fractional anisotropy (FA) is a metric derived from dMRI quantifying the degree to which water diffusion is directionally restricted in a voxel. FA values, ranging from 0 to 1, reflect the integrity of white matter, with higher FA suggesting greater structural coherence and organization (Jones et al., 2013; Kochunov et al., 2012). This technique has become a valuable tool in psychiatry research (Podwalski et al., 2021; Shizukuishi et al., 2013; White et al., 2008). Multiple studies suggest that compromised white matter integrity and associated disruptions in neural connectivity are implicated in the development and progression of both BD and DD (Duan et al., 2021; Koshiyama et al., 2020; Manelis et al., 2021; Wise et al., 2016; Xu et al., 2024). Furthermore, these neural alterations may be related to immunological and inflammatory markers, thus underscoring the complex biological mechanisms underlying mood disorders (Aronica et al., 2022).

Compared to healthy controls (HC), both BD and DD demonstrated reduced white matter integrity in key tracts, including the uncinate fasciculus (UF), the superior longitudinal fasciculus (SLF), the inferior fronto-occipital fasciculus, and the corpus callosum (CC) (Favre et al., 2019; Luttenbacher et al., 2022; Vai et al., 2020; Xu et al., 2024). Research comparing the two disorders suggests that BD may involve more pronounced white matter alterations, particularly affecting the arcuate fasciculus (AF) (Manelis et al., 2021), the UF (Fan et al., 2024), the SLF (Repple et al., 2017; Thiel et al., 2023), and the CC (Masuda et al., 2020).

Mood disorders are often linked to impaired cognitive functioning and emotion dysregulation (Alloy et al., 2006; Breukelaar et al., 2020; Korgaonkar et al., 2019; Lima et al., 2018; Manelis et al., 2019, 2022; McTeague et al., 2017; Townsend and Altshuler, 2012). One important goal of clinical neuroimaging is to understand the relationship between structural brain differences and the manifestation of cognitive and clinical symptoms. While some studies suggest that white matter integrity is independent of current symptom severity (Thiel et al., 2023), others have identified associations between white matter abnormalities and specific symptom profiles in BD and DD. For example, in DD, higher anhedonia was associated with lower FA in the CC, cingulum, and UF (Dillon et al., 2018), and longer illness duration was associated with lower FA in the CC (Zhao et al., 2021). In BD, more severe depression and anxiety symptoms were associated with lower FA in the forceps minor and anterior cingulum bundle (Santos et al., 2022a), more severe mania symptoms were related to FA in the anterior thalamic radiation and the cingulum-angular bundle (Mamah et al., 2019), and higher anxiety was associated with lower FA in the UF (Fan et al., 2024). Taken together, these findings suggest that disruptions in white matter connectivity may account for the unique clinical characteristics of BD and DD, potentially contributing to distinct presentations of mood symptoms and cognitive deficits between the two disorders.

Only a small number of neuroimaging studies have attempted to use white matter integrity measures to predict future symptom trajectories in BD and DD. A clearer understanding of these associations could inform more precise prevention, and intervention approaches for affected individuals. Recent longitudinal studies have made a promising step in that direction by revealing that FA in the right UF predicted the onset of BD over six years (Li et al., 2021), and lower FA in the right UF and cingulum predicted worsening of subthreshold hypomania in non-BD individuals over six months (Santos et al., 2022b).

Although the classical voxel-based approach to dMRI data analysis successfully identified white matter abnormalities in mood disorders, it has been criticized for using a single tensor model that is unable to accurately characterize crossing or branching fibers, thus making it difficult to differentiate between fiber crossing, myelin loss, axonal damage, and other contributing factors (Jeurissen et al., 2013; Jones and Cercignani, 2010). These methodological constraints may contribute to inconsistencies in findings when comparing white matter tract integrity in BD versus DD. Newer techniques, such as fixel-based analysis (FBA), offer an improved approach to modelling the fiber orientation distribution in complex regions with crossing fibers (Dhollander et al., 2021; Raffelt et al., 2017, 2015; Tournier et al., 2019). FBA computes measures of fiber density (FD), fiber cross-section (FC), and fiber density cross-section (FDC) in specific pathways, providing more detailed information about changes in the microstructural properties of white matter tracts. For example, reduced FC may indicate atrophy, compression, or other structural changes, while reduced FD could be linked to pathological changes in axons including axonal degeneration (Dhollander et al., 2021; Raffelt et al., 2017).

Currently, only one study has employed the FBA approach for dMRI data in the context of mood disorders (Lyon et al., 2019). This study revealed that compared to HC, individuals with DD demonstrated lower FC in the anterior limb of the internal capsule and lower FD in the CC. Importantly, depression remission at an 8-week follow-up was predicted by higher FC in the tapetum (Lyon et al., 2019), thus suggesting the potential utility of FBA measures in predicting future clinical outcomes.

No studies to date have directly compared FBA measures between individuals with BD and DD. Our study aims to fill this gap by examining fiber-specific measures of white matter integrity in individuals with BD, DD, and HC. Furthermore, we will prospectively assess dimensional symptoms of depression and mania over a six-month period following the dMRI scan. A spectrum or dimensional approach to psychopathology proposes that mental disorders exist on a continuum rather than as discrete categories (Cuthbert, 2014; Cuthbert and Insel, 2013; Insel et al., 2010). While mania and hypomania are distinct features of BD, subthreshold manifestations of hypo/mania may also occur in DD (Cassano et al., 2004; Hirschfeld et al., 2003).

Our previous research that employed spectrum measures of depression and mania in BD and DD revealed an interplay between these measures in the right middle longitudinal fasciculus and the right AF (Manelis et al., 2021). Understanding whether and how FBA measures predict future dimensional symptoms of depression and hypo/mania is crucial for elucidating the neurobiological mechanisms underlying symptom progression in BD and DD. Based on previous studies, we hypothesize that BD and DD will show distinct patterns of FD, FC, and FDC abnormalities compared to HC, with BD exhibiting more pronounced tract-specific abnormalities. Additionally, we anticipate that more severe future spectrum symptoms of depression and/or hypo/mania will be associated with lower FD, FC, and FDC in key white matter tracts (e.g., UF, CC).

## 2 METHODS

### 2.1 Participants

The study was approved by the University of Pittsburgh Institutional Review Board (IRB number: STUDY20060265). Participants were recruited from the community, universities, and counseling and medical centers, and they signed informed consent prior to participating. All participants were right-handed and fluent in English. Individuals with BD (Type I or Type II) and DD (major depressive (MDD) or persistent depressive (PDD) disorders) met DSM-5 criteria for their respective diagnoses. Healthy controls had no personal or family history of psychiatric disorders and were not taking psychotropic medications. Exclusion criteria included a history of head injury, metal in the body, pregnancy, claustrophobia, neurodevelopmental disorders, systemic medical illness, premorbid IQ<85 per the National Adult Reading Test (Nelson, 1982), current alcohol/drug abuse (within the previous 3 months), and Young Mania Rating Scale (YMRS) scores>10 (Young et al., 1978) at scan. Participants were also excluded from analyses due to changes in diagnosis (n=1 converted to MDD from HC, n=2 converted to BD Type II from MDD), or poor image quality (n=4).

### 2.2. Clinical assessments

All participants underwent clinical evaluation by a trained clinician using the Structured Clinical Interview for DSM-5 (First, 2015) to confirm their psychiatric diagnoses according to DSM-5 criteria. Participants’ diagnoses were further validated by a psychiatrist. Additionally, we collected data on participants’ medications, history of marijuana use within the previous 12 months, lifetime illness onset and duration, number of mood episodes, comorbid psychiatric disorders, (HDRS-25) (Hamilton, 1960), current mania symptoms using the YMRS (Young et al., 1978), and lifetime depression and hypo/mania spectrum symptomatology using the Mood Spectrum Self-Report questionnaire (MOODS-SR), which consists of 161 questions about mood symptoms experienced over at least a 3-5 day period (Dell’Osso et al., 2002). We also calculated a total psychotropic medication load (Hassel et al., 2008; Manelis et al., 2016), with a greater load corresponding to a higher number and doses of medications.

All participants scanned at baseline were invited to attend a 6-month follow-up appointment, which included a clinical evaluation. During this follow-up, the participants were asked to rate their spectrum depression and mania symptoms that they had experienced between the baseline scan and the 6-month appointment, using the MOODS-SR questionnaire. The MRI and behavioral data collected at the follow-up appointment will be reported elsewhere.

### 2.3 Neuroimaging data acquisition

Diffusion weighted images were collected at the University of Pittsburgh/UPMC Magnetic Resonance Research Center using a 3T Siemens Prisma scanner with a 64-channel receiver head coil. The images were acquired in the anterior-to-posterior direction with the following parameters: 92 slices, TR = 3230ms, TE=89.2ms, voxel size =1.5 mm^3^, TA=5:37, MB = 4, FOV=210mm, flip angle=78°, 46 volumes Swith *b* = 1500 s/mm^2^, 46 volumes with *b* = 3000 s/mm^2^, 7 volumes with *b* = 0 s/mm^2^. We also collected an image with 8 *b* = 0 s/mm^2^ in the posterior-to-anterior direction to apply it for geometric distortion. Unfortunately, 20% of those images were collected incorrectly due to a technical error and therefore, are not used in the analyses. Field maps were collected in the AP and PA directions using the spin echo sequence (TR=8000, resolution=2×2×2mm, FOV=210, TE=66ms, flip angle=90°, 72 slices). High-resolution T1w images were collected using the MPRAGE sequence with TR=2400ms, resolution=0.8×0.8×0.8mm, 208 slices, FOV=256, TE=2.22ms, flip angle=8°. High-resolution T2w images were collected using TR=3200ms, resolution=0.8×0.8×0.8mm, 208 slices, FOV=256, TE=563ms. The DICOM image files were named according to the ReproIn convention (Visconti di Oleggio Castello et al., 2020) and converted to BIDS dataset using *heudiconv* (Halchenko et al., 2019) and *dcm2niix* (Li et al., 2016).

### 2.4 Data analyses

#### 2.4.1 Clinical data analysis

Demographic and clinical variables for individuals in the BD, DD, and HC groups were compared using ANOVA, t-tests, and chi-square tests (whichever was appropriate) in R (https://www.r-project.org/).

#### 2.4.2 Neuroimaging data analysis

The dMRI data from 163 individuals were analyzed using MRtrix3 (Tournier et al., 2019) using the pipeline described in the MRTrix3 documentation and the B.A.T.M.A.N. tutorial (Tahedl, 2018). The data were preprocessed using denoising (Veraart et al., 2016b, 2016a), Gibbs ringing artifact removal (Kellner et al., 2016), as well as motion, distortion (Andersson et al., 2003; Andersson and Sotiropoulos, 2016; Smith et al., 2004), and bias (Tustison et al., 2010) correction. We utilized synb0-DISCO for distortion correction (Schilling et al., 2020, 2019).

After preprocessing, the response functions for white matter (WM), grey matter (GM), and cerebrospinal fluid (CSF) (Dhollander et al., 2016) were estimated to perform constrained spherical deconvolution (CSD) (Tournier et al., 2007, 2004). All individual subject brain masks were computed using the *bet* command in FSL (Smith, 2002) before estimating Fiber Orientation Distributions (FODs). FODs were estimated via multi-shell multi-tissue CSD (Jeurissen et al., 2014) using *dwi2fod* function in MRtrix to estimate the orientation of the fibers crossing each voxel (Tournier et al., 2007, 2004). The dMRI images were not upsampled before computing FODs due to the already high original resolution (an isotropic voxel size for original images was 1.5mm^3^). Joint bias field correction and global intensity normalization of the multi-tissue compartment parameters were performed using the *mtnormalise* command for all subjects (Dhollander et al., 2021; Raffelt et al., 2017). A study-specific population template was created by averaging the FOD data from all 163 participants. All individuals subject FOD images were nonlinearly registered to the FOD template (Raffelt et al., 2017). The template mask was computed as the intersection of all warped masks in template space, ensuring that only voxels with data from all subjects were included. A fixel mask was then generated from the FOD template using *fod2fixel* function, defining the fixels for statistical analysis. After that, a fixel-based analysis was conducted by segmenting and reorienting participants’ fixels to the template and calculating apparent fiber density (FD), fiber cross-section (log(FC)), and fiber density and cross-section combined (FDC) metrics for each participant. TractSeg (Wasserthal et al., 2019, 2018) was used to directly segment 72 white matter tracts from the peaks of the spherical harmonic function at each voxel. Probabilistic bundle-specific tractography was then performed on the tract orientation maps. Next, a fixel-fixel connectivity matrix was generated based on the tractogram, followed by statistical analysis of FD, FC, and FDC.

Statistical analyses were conducted using the *fixelcfestats* tool in the MRtrix3. An F-test was employed to compare BD, DD, and HC groups within each tract using smoothed data within a general linear model framework with 3000 permutations and family-wise error-corrected (FEW) p-values < 0.05. Following this, we performed linear regression analyses to examine the relationship between white matter integrity and future symptom trajectories. Specifically, we tested whether the interaction between diagnostic group (BD vs. DD) and fixel-based metrics (e.g., FD values) predicted dimensional measures of depression or hypo/mania over the six-month follow-up period. These analyses were restricted to tracts identified in the primary group comparison. Covariates included age, sex, IQ, and lifetime spectrum measures of depression or mania to account for baseline differences. Additionally, exploratory analyses were conducted to assess potential confounding factors. We examined the effects of psychotropic medication use (medicated vs. unmedicated and medication load), history of marijuana use within the past 12 months, and illness duration on FBA metrics that showed significant group differences in the primary analysis.

## 3. RESULTS

### 3.1 Clinical results

Of the 170 individuals recruited for the study, dMRI data from 163 participants were included in the analyses. Demographic and clinical characteristics are presented in Table 1. The BD, DD, and HC groups did not differ in sex composition. However, individuals with BD were younger and had slightly higher IQ scores than HC. No significant differences were found between the BD and DD groups in terms of age of illness onset, illness duration, number of individuals with comorbid psychiatric disorders, or YMRS scores. However, individuals with BD reported significantly more severe lifetime spectral symptoms of mania and general psychopathology compared to those with DD (*p* < 0.001 for all comparisons). Additionally, a greater proportion of individuals with BD were taking psychotropic medications compared to those with DD (*p* = 0.03). Among those taking psychotropic medications, individuals with BD had a higher medication load (*p* = 0.001).

**Table 1.**
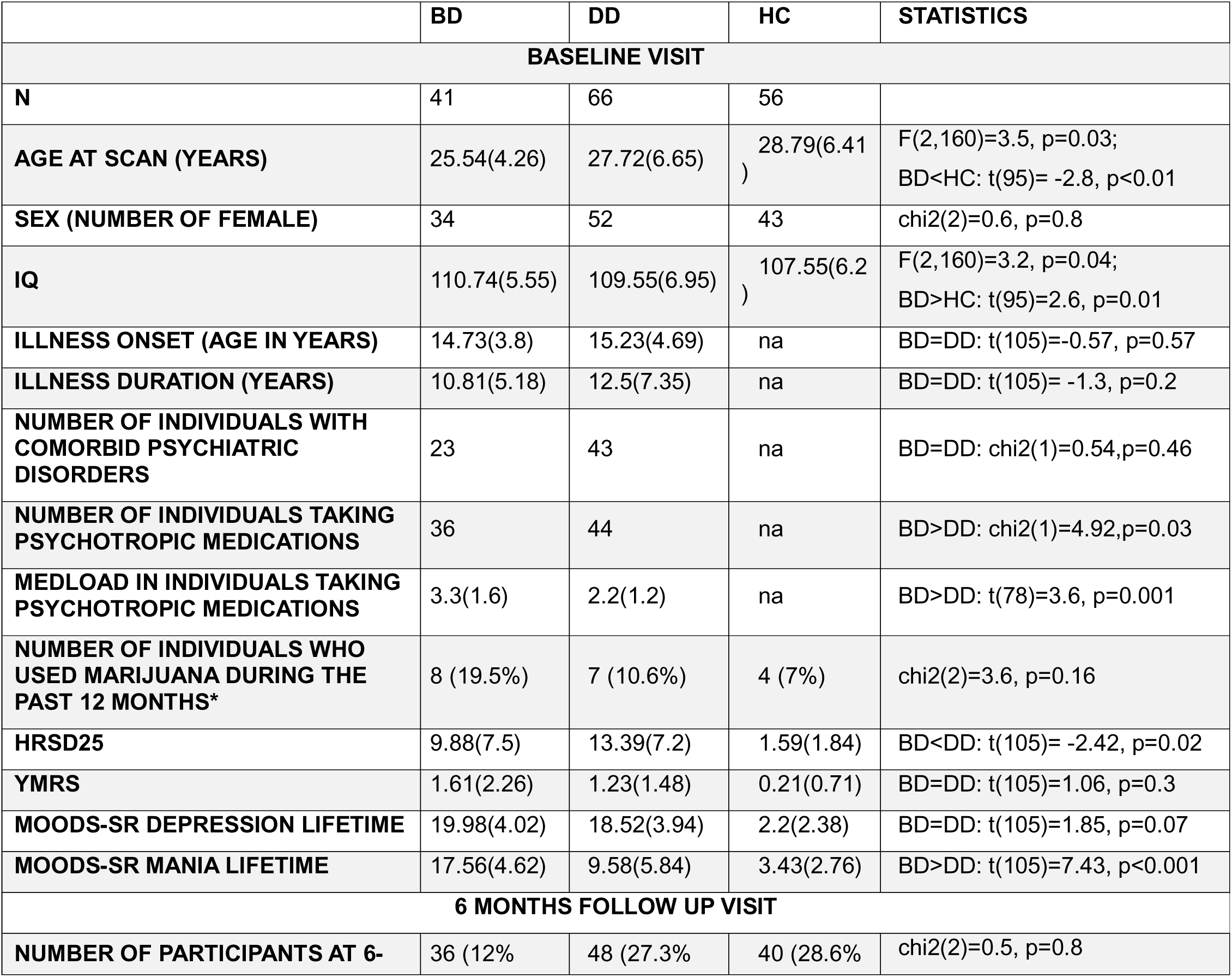
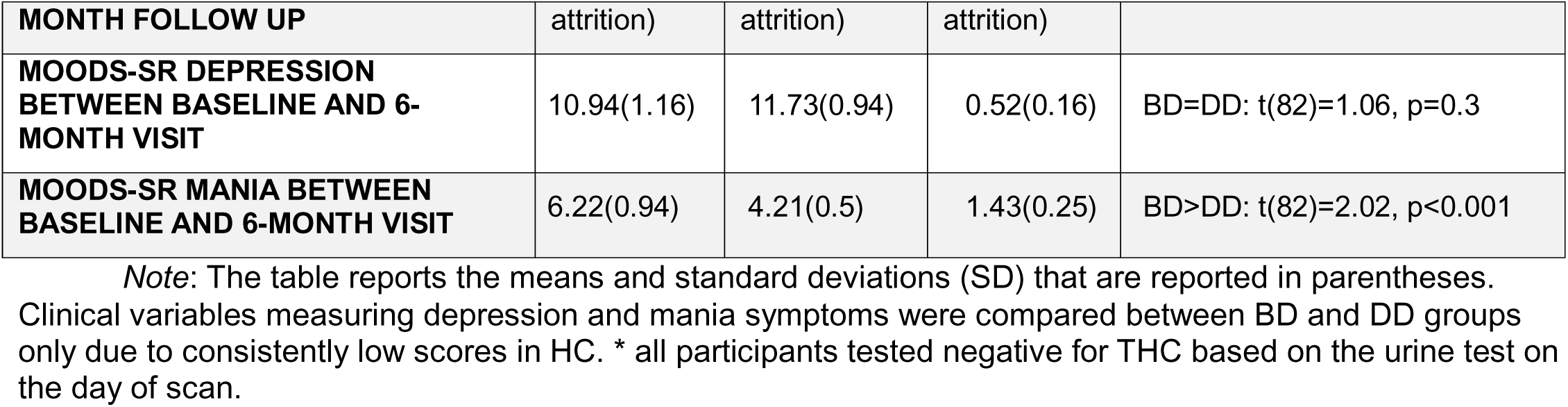
Demographic and clinical characteristics.

|  | BD | DD | HC | STATISTICS |
| --- | --- | --- | --- | --- |
| **BASELINE VISIT** |  |  |  |  |
| **N** | 41 | 66 | 56 |  |
| **AGE AT SCAN (YEARS)** | 25.54(4.26) | 27.72(6.65) | 28.79(6.41) | $F(2,160)=3.5, p=0.03$;<br>BD<HC: $t(95)= -2.8, p<0.01$ |
| **SEX (NUMBER OF FEMALE)** | 34 | 52 | 43 | $\chi^2(2)=0.6, p=0.8$ |
| **IQ** | 110.74(5.55) | 109.55(6.95) | 107.55(6.2) | $F(2,160)=3.2, p=0.04$;<br>BD>HC: $t(95)=2.6, p=0.01$ |
| **ILLNESS ONSET (AGE IN YEARS)** | 14.73(3.8) | 15.23(4.69) | na | BD=DD: $t(105)=-0.57, p=0.57$ |
| **ILLNESS DURATION (YEARS)** | 10.81(5.18) | 12.5(7.35) | na | BD=DD: $t(105)= -1.3, p=0.2$ |
| **NUMBER OF INDIVIDUALS WITH COMORBID PSYCHIATRIC DISORDERS** | 23 | 43 | na | BD=DD: $\chi^2(1)=0.54, p=0.46$ |
| **NUMBER OF INDIVIDUALS TAKING PSYCHOTROPIC MEDICATIONS** | 36 | 44 | na | BD>DD: $\chi^2(1)=4.92, p=0.03$ |
| **MEDLOAD IN INDIVIDUALS TAKING PSYCHOTROPIC MEDICATIONS** | 3.3(1.6) | 2.2(1.2) | na | BD>DD: $t(78)=3.6, p=0.001$ |
| **NUMBER OF INDIVIDUALS WHO USED MARIJUANA DURING THE PAST 12 MONTHS\*** | 8 (19.5%) | 7 (10.6%) | 4 (7%) | $\chi^2(2)=3.6, p=0.16$ |
| **HRSD25** | 9.88(7.5) | 13.39(7.2) | 1.59(1.84) | BD<DD: $t(105)= -2.42, p=0.02$ |
| **YMRS** | 1.61(2.26) | 1.23(1.48) | 0.21(0.71) | BD=DD: $t(105)=1.06, p=0.3$ |
| **MOODS-SR DEPRESSION LIFETIME** | 19.98(4.02) | 18.52(3.94) | 2.2(2.38) | BD=DD: $t(105)=1.85, p=0.07$ |
| **MOODS-SR MANIA LIFETIME** | 17.56(4.62) | 9.58(5.84) | 3.43(2.76) | BD>DD: $t(105)=7.43, p<0.001$ |
| **6 MONTHS FOLLOW UP VISIT** |  |  |  |  |
| **NUMBER OF PARTICIPANTS AT 6-** | 36 (12%) | 48 (27.3%) | 40 (28.6%) | $\chi^2(2)=0.5, p=0.8$ |

| MONTH FOLLOW UP | attrition) | attrition) | attrition) |  |
| --- | --- | --- | --- | --- |
| **MOODS-SR DEPRESSION BETWEEN BASELINE AND 6-MONTH VISIT** | 10.94(1.16) | 11.73(0.94) | 0.52(0.16) | BD=DD: $t(82)=1.06$, $p=0.3$ |
| **MOODS-SR MANIA BETWEEN BASELINE AND 6-MONTH VISIT** | 6.22(0.94) | 4.21(0.5) | 1.43(0.25) | BD>DD: $t(82)=2.02$, $p<0.001$ |

*Note:* The table reports the means and standard deviations (SD) that are reported in parentheses. Clinical variables measuring depression and mania symptoms were compared between BD and DD groups only due to consistently low scores in HC. \* all participants tested negative for THC based on the urine test on the day of scan.

Regarding symptom trajectories between baseline and the 6-month follow-up, individuals with BD exhibited greater spectrum hypo/mania symptom severity compared to those with DD (*t*(82)=2.02, *p*=0.047; BD: *M*=6.2±5.7, DD: *M*=4.2±3.5). However, no significant group differences were observed in spectrum depression severity during this period (*t*(82)= −0.5, *p*=0.6; BD: *M*=10.9±6.9, DD: *M*=11.7±6.6).

### 3.2 Tractography results

#### 3.2.1 Group comparison

Fixel-based analysis (FBA) revealed a significant main effect of diagnosis on fiber density (FD) in 9 tracts (FWER-p<0.05). These tracts included the bilateral arcuate fascicle (AF), the splenium of corpus callosum (CC 7), the right uncinate fascicle (UF), the right superior longitudinal fascicles II and III (SLF II and SLF III), the left optic radiation (OR), the left thalamo-occipital (T-OCC), and the left striato-occipital (ST-OCC) tracts (Figure 1). No significant effect of diagnosis was observed on FC or FDC measures. To further explore the effects in each of the identified tracts, we extracted FD values from the significant fixels and computed pairwise group-level contrasts using the estimates from linear regression models (one-way ANOCOVA with age, sex, and IQ as covariates), adjusting for multiple comparisons using the Tukey HSD post-hoc test. The results are reported in Table 2. Age, sex, and IQ did not affect FD values in any of the 9 tracts.

**Figure 1.**
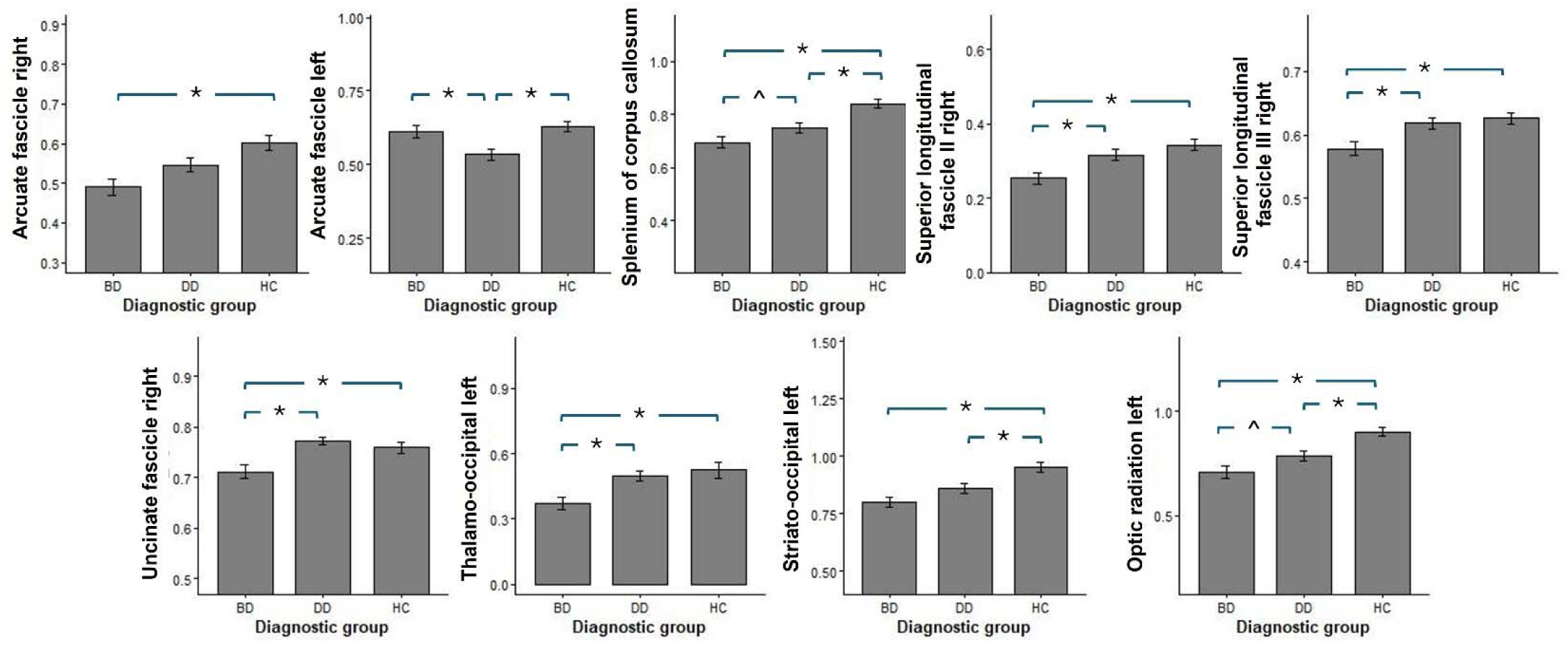
Fiber density (FD) alterations in individuals with bipolar and depressive disorders relative to HC. Asterisks (*) denote statistically significant differences between the groups, and caret symbols (^) denotes marginally significant differences.

**Table 2.** The pairwise group-level contrasts computed using the estimates from the linear regression models and adjusted for multiple comparisons using Tukey HSD post-hoc test.

| Tract | Analysis of contrasts (df=157), p-corrected using Tukey HSD post-hoc test |
| --- | --- |
| Arcuate fascicle (AF) left | HC=BD, HC>DD ($t=3.6$, $p=0.001$), BD>DD ($t=2.4$, $p=0.04$) |
| Arcuate fascicle (AF) right | HC>BD ($t=3.5$, $p=0.002$), HC=DD, BD=DD |
| Splenium of corpus callosum (CC 7) | HC>BD ($t=5.3$, $p<0.001$), HC>DD ($t=3.8$, $p<0.001$), BD<DD ($t=-2.2$, $p=0.08$)^ |
| Superior longitudinal fascicle II (SLF II) right (association) | HC>BD ($t=4.0$, $p<0.001$), HC=DD, BD<DD ($t=-2.9$, $p=0.01$) |
| Superior longitudinal fascicle III (SLF III) right (association) | HC>BD ($t=3.2$, $p=0.004$), HC=DD, BD<DD ($t=-2.9$, $p=0.01$) |
| Uncinate fascicle (UF) right | HC>BD ($t=3.3$, $p=0.003$), HC=DD, BD<DD ($t=-2.9$, $p=0.01$) |
| Thalamo-occipital (T-OCC) left | HC>BD ($t=3.3$, $p=0.003$), HC=DD, BD<DD ($t=-2.9$, $p=0.01$) |
| Striato-occipital (ST-OCC) left | HC>BD ($t=4.1$, $p<0.001$), HC>DD ($t=3.0$, $p=0.01$), BD=DD |
| Optic radiation (OR) left | HC>BD ($t=4.9$, $p<0.001$), HC>DD ($t=3.4$, $p=0.002$), BD<DD ($t=-2.1$, $p=0.09$)^ |

*Note:* ^ - marginally significant

#### 3.2.2 Predicting future spectrum symptoms of depression and hypo/mania from FD in individuals with BD and DD

Analyses predicting the severity of spectrum depression over the 6-month period based on the interactions between the BD/DD group and FD in the tracts identified in the primary analysis revealed significant interaction effects in the splenium of corpus callosum (CC 7-by-group interaction: F(1,76)=4.38, p=0.04), the left striato-occipital (ST-OCC-by-group interaction: F(1,76)=4.35, p=0.04), and the right superior longitudinal fascicle II (SLF II-by-group interaction, F(1,76)=13.4, p=0.0005) tracts. Interestingly, in individuals with BD, more severe spectrum symptoms of depression were predicted by lower FD in CC 7 and the left ST-OCC but by higher FD in the right SLF II. The opposite pattern was observed in the DD group (Figure 2A).

**Figure 2.**
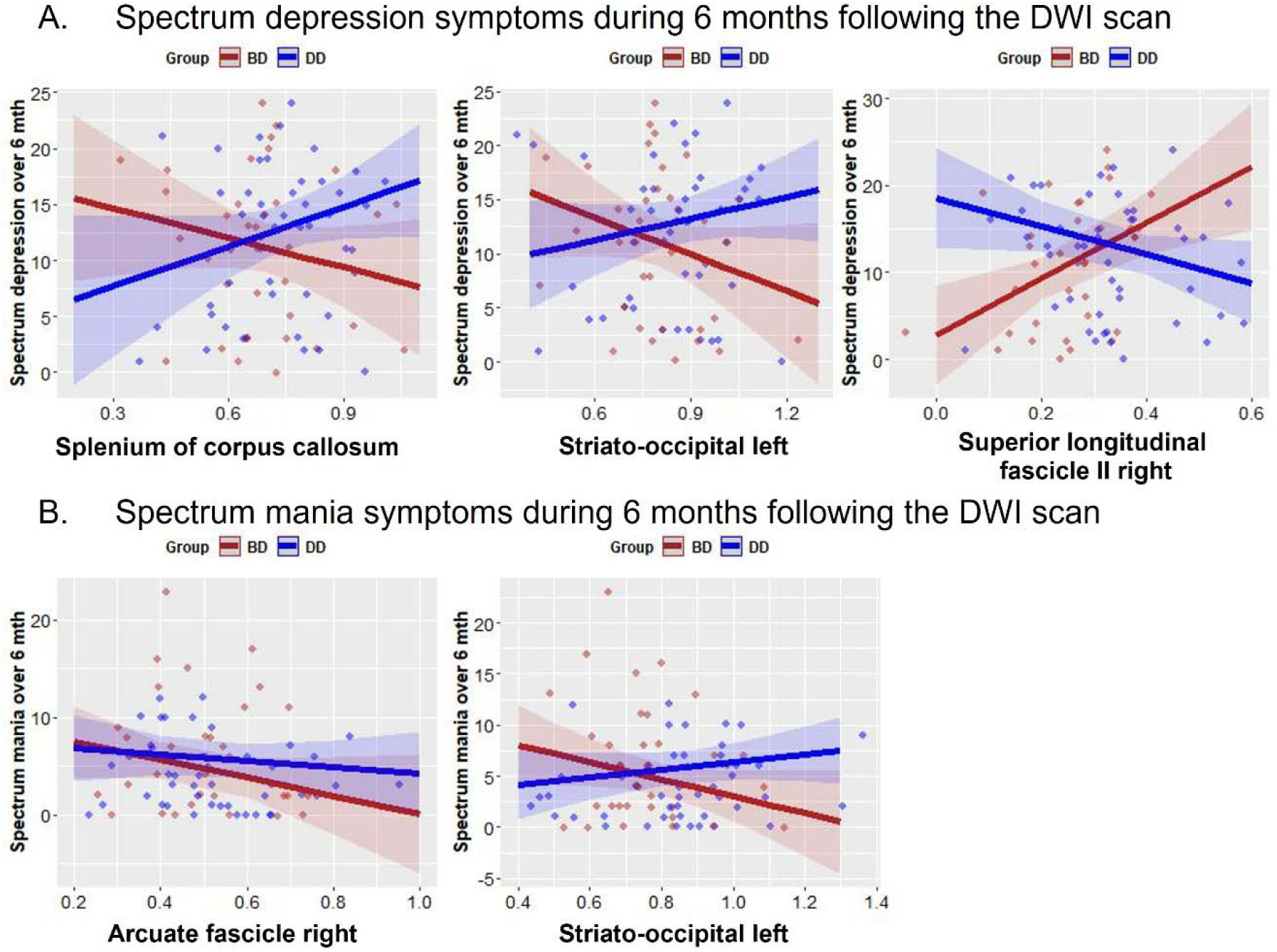
Predicting spectrum depression and hypomania from the interaction between the diagnostic group and FD values.

**Figure 3.**
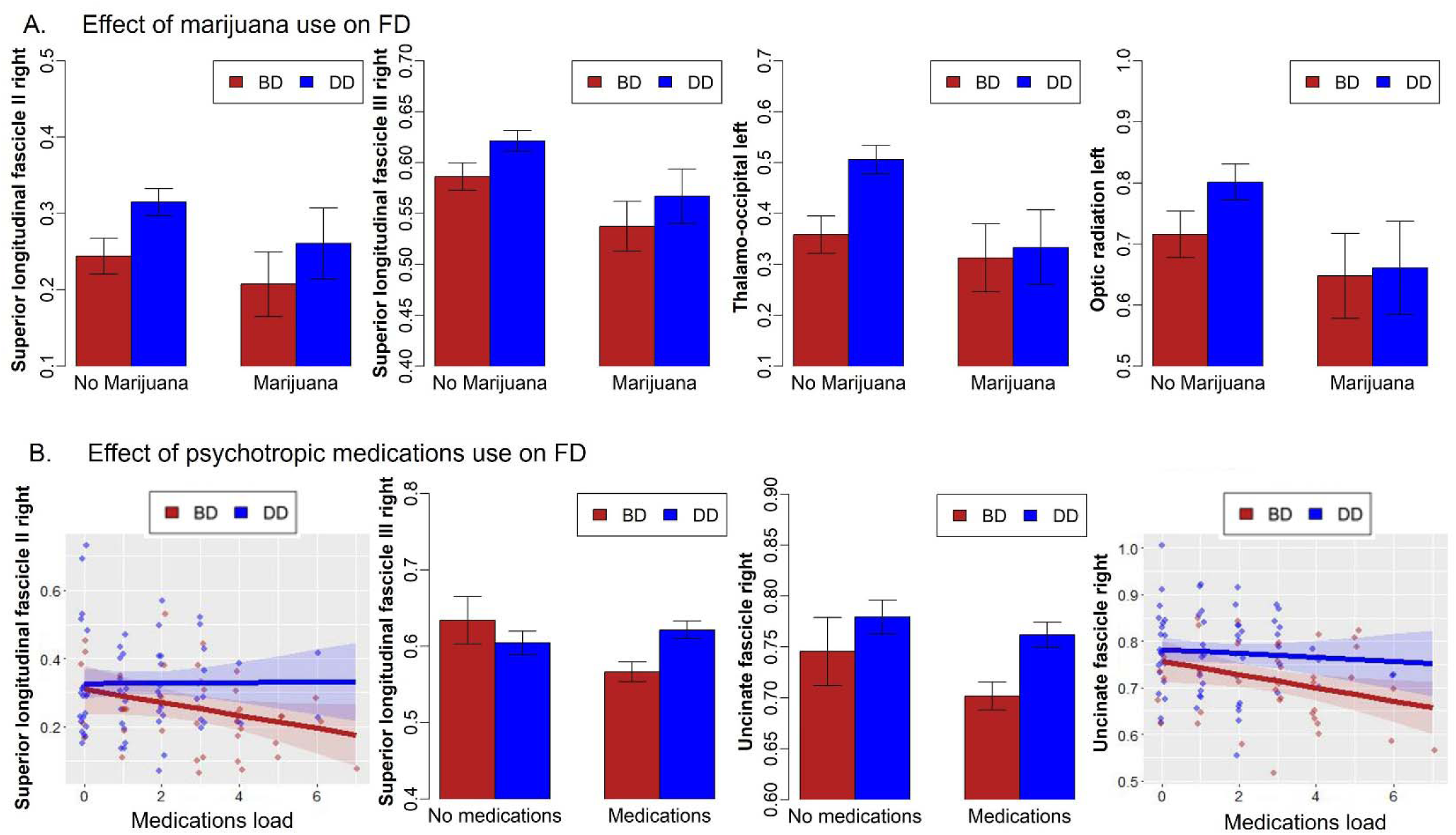
The effects of marijuana and psychotropic medications on FD values in individuals with BD and DD.

More severe spectrum mania symptoms were observed across diagnoses in individuals who had lower FD in the right AF (main effect of FD: F(1,76)=4.8, p=0.03). More severe spectrum symptoms of mania were observed in those with BD who had lower FD in the left ST-OCC and those with DD who had higher FD in the same tract (ST-OCC-by-group interaction: F(1,76)=4.46, p=0.037).

#### 3.3.3 Exploratory analyses

This exploratory analysis investigated the effect of interaction between diagnostic group with illness onset and duration as well as medications and marijuana use on FD in the tracts identified in the main analyses, controlling for age, sex, and IQ. The main effect of illness duration was found in the right UF with longer duration associated with higher FD (*F*(1,100)=5.3, *p*=0.02; *t*=2.2, *p*=0.02). There were no significant main or interaction effects of illness onset on FD in any of the 9 tracts.

Marijuana use in the past 12 months was linked to lower FD in the left OR (*F*(1,100)=4.3, *p*=0.04), right SLF II (*F*(1,100)=4.4, *p*=0.04) and SLF III (*F*(1,100)=8.2, *p*=0.005), and left T-OCC (*F*(1,100)=7.5, *p*=0.008), independent of BD or DD diagnosis. Medication effects were observed in the right SLF II, SLF III, and UF.

In the right SLF II, higher medload was associated with lower FD (*F*(1,100)=4.8, *p*=0.03), regardless of diagnosis. In the right SLF III, an interaction between medication use and diagnosis (*F*(1,100)=5.1, *p*=0.026) indicated a stronger effect of medication use in BD (*t*=2.26, *p*=0.03). In the right UF, both medication use (*F*(1,100)=5.24, *p*=0.0242) and medload (*F*(1,100)=12.44, *p*=0.0006) significantly affected FD, with lower FD observed in individuals on higher medication doses.

## 4. DISCUSSION

In this study, we conducted a whole brain fixel-based analysis (FBA) (Dhollander et al., 2021; Raffelt et al., 2017, 2015; Tournier et al., 2019) to compare white matter fiber density (FD), fiber cross-section (FC), and fiber density-cross-section (FDC) among individuals with BD, DD, and HC. Our findings revealed significant differences in FD, but not FC or FDC, across multiple white matter tracts. Compared to HC, individuals with BD exhibited lower FD in the right arcuate fasciculus (AF), splenium of the corpus callosum (CC), right superior longitudinal fasciculus II and III (SLF II and SLF III), right uncinate fasciculus (UF), left temporo-occipital (T-OCC) tract, left superior temporal-occipital (ST-OCC) tract, and left optic radiation (OR). Similarly, individuals with DD showed lower FD in the left AF, splenium of the CC, left ST-OCC, and left OR compared to HC. Direct comparisons between BD and DD indicated lower FD in BD within the right SLF II and III, right UF, and left T-OCC, whereas DD exhibited lower FD in the left AF compared to BD. These findings expand upon previous research that reported lower FD in the CC of individuals with DD compared to HC (Lyon et al., 2019).

FD reflects white matter microstructure, with lower values indicating reduced intra-axonal volume, potentially related to neurodegeneration or axonal loss (Dhollander et al., 2021; Genc et al., 2020; Raffelt et al., 2017, 2012). Our findings of reduced FD in the AF, UF, CC, and SLF II and III align with prior studies using FA to assess white matter integrity (Fan et al., 2024; Manelis et al., 2021; Repple et al., 2017; Thiel et al., 2023; Yip et al., 2013). The AF tract connects frontal, parietal, and temporal lobes, including Broca’s and Wernicke’s areas, playing a key role in language processing (Bain et al., 2019; Catani and Mesulam, 2008; Fernández-Miranda et al., 2015; Radwan et al., 2022). Previous studies have linked AF abnormalities to mood disorders (Manelis et al., 2021; Sun et al., 2017) and found associations with mood episode frequency in BD (Raucher-Chéné et al., 2017).

The UF links the ventral, medial and orbital frontal lobes with rostral temporal lobes and is critical for emotion regulation and social cognition (Radwan et al., 2022; Von Der Heide et al., 2013). The observation of lower FD in this region in BD and DD compared to HC aligns with previous findings (Foley et al., 2018; Santos et al., 2022b; Xu et al., 2022) and may contribute to the emotion dysregulation characterizing these disorders.

The SLF is an association fiber tract that connects the frontal and parietal lobes including the temporoparietal junction area (Wang et al., 2016). The right SLF II is implicated in visuospatial processing, including spatial attention and orientation (De Schotten et al., 2011), while the right SLF III plays a role in spatial information processing (Wang et al., 2016). The CC, a commissural tract connecting the right and left hemispheres (Radwan et al., 2022), is essential for transferring information between hemispheres to support optimal visuomotor integration and cognitive processes (Schulte and Müller-Oehring, 2010). Our findings of lower FD in individuals with mood disorders compared to HC align with previous research(Xu et al., 2024; Yang et al., 2019), suggesting disrupted interhemispheric communication, which may underlie the aberrant visuomotor processing observed in these individuals (Hu et al., 2024; Tsai et al., 2023). Additionally, our observation of reduced FD in the splenium of the CC is consistent with prior studies reporting lower myelin content in this region in individuals with DD compared to HC (Williams et al., 2019). Together, reduced FD in the SLF and CC may explain visuospatial and visuomotor deficits previously observed in mood disorders (Gallagher et al., 2015; Nelson and Shankman, 2016; Pan et al., 2011).

To our knowledge, our study is the first to report reduced FD in the T-OCC, ST-OCC, and OR in individuals with BD and DD compared to HC. These projection tracts connect the occipital lobe with the thalamus, striatum, and lateral geniculate nucleus (LGN) of the thalamus, respectively. The OR is involved in visual information transmission and visual working memory (Wang et al., 2024), while the T-OCC contributes to visual perception (Joo et al., 2018), visual attention (Menegaux et al., 2020), and cognitive performance (Byun et al., 2020). Though less studied, the ST-OCC likely coordinates visual perception with motor responses, reward processing, and decision-making. Lower FD in T-OCC, ST-OCC, and OR tracts has been linked to lower semantic fluency (Egorova-Brumley et al., 2022), highlighting their role in semantic memory and the integration of visual and language information. Our findings suggest that FD reductions in these pathways may underlie verbal fluency and information-processing deficits in mood disorders (Alloy et al., 2012, 2006; Raucher-Chéné et al., 2017), potentially impairing visuomotor planning, visual attention, and decision-making.

### Using FD to predict future symptom severity

Beyond examining the impact of diagnosis on white matter microstructure, our study explored whether FD could predict spectrum symptoms of depression and hypo/mania over the 6 months following the scan. In BD, more severe depressive symptoms were associated with lower FD in the splenium of the CC and left ST-OCC but higher FD in the right SLF-II. Conversely, in DD, greater depression severity was linked to higher FD in the CC7 and left ST-OCC and lower FD in the right SLF-II. For hypo/mania, lower FD in the right AF predicted greater symptom severity across mood disorder groups. Additionally, FD in the left ST-OCC was associated with hypo/mania severity, with BD showing more pronounced symptoms at lower FD levels, while DD exhibited greater severity at higher FD levels. It is noteworthy that the UF was not among the tracts linked to future mood symptoms, contrary to prior longitudinal studies where FA in the UF predicted subthreshold hypomania severity (Santos et al., 2022b) and BD onset (Li et al., 2021) in individuals not yet diagnosed with BD.

These findings align with dimensional models of psychopathology (Cuthbert, 2014; Cuthbert and Insel, 2013; Insel et al., 2010) while emphasizing that shared neural substrates may contribute differently to symptom presentation depending on diagnosis. White matter alterations in BD have been linked to neuroinflammation (Benedetti et al., 2020), suggesting that reduced FD may reflect neuroinflammatory processes underlying spectrum symptoms of depression and mania. The relationship between symptom severity and higher FD is more complex and challenging to interpret. On the one hand, elevated FD may reflect compensatory neuroplastic changes in response to mood dysregulation in BD and DD. Alternatively, it may reflect disruptions in normal white matter organization, affecting cognitive, emotional, and sensorimotor coordination. Further research is needed to fully elucidate the complex relationship between white matter microstructure and mood disorder pathology (Xu et al., 2024).

### The effect of medication and marijuana use on FD

Our exploratory analysis revealed that longer illness duration was associated with greater FD in the right UF, suggesting a potential compensatory reorganization in response to chronic illness. This contrasts with previous studies reporting a negative correlation between illness duration and FA in the CC (Yang et al., 2021) or no relationship between illness duration and FA at all (Manelis et al., 2021), possibly due to the greater specificity of FBA compared to conventional diffusion-tensor analysis. Although previous studies observed the effect of age on FD in the CC, SLF, and some other tracts (Choy et al., 2020), no effects of age, sex or IQ were identified in our study. One explanation for this is that our sample consisted of younger adults not yet experiencing degeneration.

Although some of the previous studies did not implicate medication use (aside from lithium) in white matter tract integrity in BD (Benedetti et al., 2011), we found that medication use was associated with lower FD in the right SLF II, SLF III, and UF, with a stronger effect in BD than DD for the SLF III, while the SLF II and UF effects were diagnosis-independent. Although the causality of this relationship cannot be inferred, our findings raise questions about whether group differences in FD reflect disease pathophysiology (as more severe symptoms may require high medication load), treatment effects, or treatment failures. Unfortunately, the relatively small sample of individuals with BD limited our ability to perform analyses by medication class.

Our study is the first to examine marijuana use history in relation to FD differences in mood disorders. Although all participants tested negative for tetrahydrocannabinol (THC) at the time of the scan, prior use within the past year was associated with reduced FD in the left OR, right SLF II, right SLF III, and left T-OCC, independent of diagnosis. These findings parallel the results of a recent meta-analysis linking marijuana use to lower white matter integrity in the AF, SLF, and CC tracts (Robinson et al., 2023) and those reporting its association with worsening mood disorder symptoms and functioning (Tourjman et al., 2023). Together, these results suggest potential long-term effects of substance use on white matter microstructure and underscore the importance of considering substance use in neuroimaging-based predictive models for symptom progression.

### Limitations and future directions

One strength of this study is its longitudinal design enabling inferences about the causal role of FD in predicting future subthreshold symptoms of depression and hypo/mania. However, the high attrition rate ranging from 12% to 29%, represents a limitation that may restrict longitudinal interpretations. Although FD predicted future spectrum symptoms, replication in larger samples is needed to improve generalizability. While we accounted for age, sex, and IQ, other factors like social determinants of health, lifestyle, and genetics may also impact white matter. The link between FD and medication use/dosage is complex, as medication effects are hard to separate from diagnosis and illness severity. Another limitation is that marijuana use data relied on self-reporting without quantifying exposure.

Future research should track longitudinal FD changes along with symptoms and treatment. For example, the diagnostic specificity of the splenium of CC 7 and the right SLF II warrants further investigation as potential biomarkers to differentiate BD and DD. Our work advances the understanding of mood disorders by identifying transdiagnostic and diagnosis-specific white matter signatures, supporting a model that integrates categorical and dimensional approaches to psychopathology.

Further longitudinal studies are needed to clarify how white matter integrity predicts the course and prognosis of mood disorders. Elucidating these brain-behavior relationships could lead to the development of neuroimaging-based biomarkers for early detection, diagnosis, and personalized treatment. For example, a recent study suggests that using tractography could benefit the DBS treatment approach for intractable depression (Chan et al., 2024). Additional research is needed to understand why using marijuana and psychotropic medications might reduce fiber density in UF and SLF tracts. Our preliminary results should be interpreted with caution because only a small number (n=5) of individuals with BD did not take psychotropic medications, and less than 10 individuals per diagnostic group reported using marijuana in the past 12 months.

### Conclusion

This study provides evidence of distinct and overlapping white matter alterations in BD and DD, with BD exhibiting more pronounced reductions in FD. Our findings also suggest that FD alterations in key tracts may predict future symptom severity, underscoring the potential utility of white matter integrity as a biomarker for mood disorder prognosis. Future longitudinal studies should prospectively evaluate the effect of treatment strategies on white matter microstructure to refine our understanding of mood disorder pathophysiology and improve clinical outcomes.

## FUNDING ACKNOWLEDGEMENTS

This work was supported by grants from the National Institute of Health R01MH114870 to A.M.

## DISCLOSURE

A.M., H.H., S.S., and S.I. declare no conflict of interest. H.A.S.: receives royalties from Wolters Kluwer, royalties and an editorial stipend from APA Press, and honorarium from Novus Medical Education.

## ACKNOWLEDGMENTS

The authors thank participants for taking part in this research study. We also thank Rachel Miceli for helping with data collection.

## AUTHOR STATEMENT

A.M. – Conceptualization, Data curation, Formal analysis, Funding acquisition, Investigation, Project administration, Supervision, Validation, Visualization, Writing – original draft, Writing – review & editing

H.H. – Data curation, Formal analysis, Methodology, Software, Validation, Visualization, Writing – review & editing

S.S. – Data curation, Investigation, Project administration, Validation, Writing – review & editing

S.I. – Conceptualization, Methodology, Validation, Writing – review & editing

H.A.S. – Conceptualization, Investigation, Supervision, Validation, Writing – review & editing.

All authors have read and approved the final version of the manuscript and agreed to be accountable for all aspects of this work.

## DATA AVAILABILITY

The data that support the findings of this study are openly available in The National Institute of Mental Health Data Archive (NDA) at https://nda.nih.gov/, reference number Collection ID: 2908.

